# From bedside to bench and back: discovery of a novel missense variant in NLRP3 causing atypical CAPS with hearing loss as the primary presentation, responsive to anti-IL-1 therapy

**DOI:** 10.1101/2023.05.05.23289337

**Authors:** Merav Birk-Bachar, Hadar Cohen, Efrat Sofrin-Drucker, Nesia Kropach-Gilad, Naama Orenstein, Gabriel Lidzbarsky, Liora Kornreich, Rotem Tal, Gil Amarilyo, Yoel Levinsky, Meirav Sokolov, Eyal Raveh, Motti Gerlic, Liora Harel

## Abstract

Cryopyrin-associated periodic syndromes (CAPS) also known as NLRP3-associated auto-inflammatory diseases, are a spectrum of rare auto-inflammatory diseases caused by gain-of-function mutations in the NLRP3 gene, resulting in inflammasome hyper-activation and dysregulated release of Interleukin-1^β^ (IL-1^β^). Many patients with CAPS develop progressive sensorineural hearing loss (SNHL) due to cochlear auto-inflammation which, in rare cases, may be the sole manifestation. This study was undertaken to establish the suspected diagnosis of CAPS in a family presenting autosomal dominant progressive/acute SNHL and a novel missense variant in the NLRP3 gene of unknown significance (NM_001079821:c.1790G>A, p.Ser597Asn). To do so, we conducted an ex vivo functional assessment of the NLRP3 inflammasome in carries (n=10) and healthy family members (n=5). The assay revealed hyper-activation of the inflammasome among carriers, supporting the hypothesis that this missense variant is a pathogenic gain-of-function mutation. Administration of anti-IL-1 therapy resulted in a substantial clinical improvement among pediatric patients, who exhibited near resolution of hearing impairment within 1-3 months of treatment. Our findings highlight the crucial role of early diagnosis and treatment of hearing loss due to hyperactivation of the inflammasome with an anti-IL-1 agent in reversing cochlear damage. Furthermore, our results suggest that high and ultrahigh frequency ranges need to be included in the auditory assessment to enable early detection of subclinical SNHL. Finally, incorporating functional inflammasome assessment as part of the clinical evaluation could establish the diagnosis in inconclusive cases.

## Introduction

The NLRP3 inflammasome[1] is an intracellular innate immune sensor expressed in immune cells, including monocytes and macrophages. Gain-of-function mutations in the NLRP3 gene lead to hyperactivation of the NLRP3 inflammasome, resulting in the inappropriate release of inflammatory cytokines, including IL-1β, and causing a spectrum of autosomal-dominant systemic autoinflammatory diseases called cryopyrin-associated periodic syndromes (CAPS) or NLRP3-associated autoinflammatory diseases (NLRP3-AIDs)[2,3]. The CAPS spectrum includes three classical clinical subtypes: neonatal-onset multisystem inflammatory disease (NOMID, MIM 607115), Muckle-Wells syndrome (MWS, MIM 191900), and familial cold autoinflammatory syndrome (FCAS, MIM 120100). The age at onset, as well as the severity of symptoms, vary within this spectrum and include recurrent fever, typical urticaria-like rash, headache, conjunctivitis, and arthralgia or arthritis.[3] Many of these patients, particularly those with the more severe phenotypes, also develop progressive sensorineural hearing loss (SNHL) due to cochlear auto-inflammation(Yamada et al., 2022(. In rare cases, SNHL may be the primary finding, and it was suggested that cochlear auto-inflammation was caused by hyper-activation of the NLRP3 inflammasome in tissue-resident macrophage/monocyte-like cells [5,6].

Inflammasomes are a group of cytosolic protein complexes located mainly in myeloid lineage cells that are formed to mediate host immune responses to microbial infection and cellular damage [7]. The NLRP3 inflammasome activation process is dependent on a two-signal sequence. In the absence of either signal, the inflammasome remains inactive. The first “priming” step, initiated by either pathogen-associated molecular pattern molecules (PAMPs), danger-associated molecular patterns (DAMPs), or pro-inflammatory cytokines, activates transcriptional factors NF-κB/AP1, resulting in the upregulation of inflammasome components as pro-caspase-1 and pro-inflammatory cytokines (i.e., pro-IL-1β and pro-IL-18). The second “activating” step, initiated by the recognition of either PAMPs or DAMPs, results in the oligomerization of inflammasome protein components [8]. Inflammasome formation then leads to auto-cleavage of caspases-1 and subsequent cleavage of gasdermin-D (GSDMD), which tends to translocate to the outer membrane to form pores and subsequently induce pyroptotic cell death. Inflammasome activation also cleaves pro-IL-1β and pro-IL-18 to their mature and active forms, which are then released through the GSDMD pores, creating a proinflammatory cascade [9,10]. In the case of NLRP3 gain-of-function mutations, the threshold for inflammasome activation may be either impaired or totally lost, resulting in inappropriate activation of the inflammasome. This hyperactivity is classically demonstrated *ex-vivo* by measuring levels of IL-1β in supernatants of PBMCs, in response to NF-kB stimulation alone (priming signal).

In this study, we present a family that initially presented with autosomal dominant SNHL and a novel variant in the NLRP3 gene of unknown significance, which did not meet CAPS classification criteria. By conducting an *ex-vivo* functional assessment of the NLRP3 inflammasome, which showed inflammasome hyperactivity, we strengthened the suspected diagnosis of CAPS. Finally, following this assessment, we applied an anti-IL-1 treatment that resulted in a rapid substantial clinical hearing improvement in young patients.

## Results

### Hearing loss as the main or sole presentation of CAPS

The proband, a two-year-old boy at the time of study initiation (III1), was initially referred to the Pediatric Rheumatology Unit due to recurrent episodes of fever and was eventually clinically diagnosed with PFAPA (periodic fever, aphthous stomatitis, pharyngitis, and adenitis). Family history revealed six family members spanning three generations who had previously been diagnosed with autosomal dominant, bilateral, progressive SNHL (Figure 1A, Table 1). The eldest (1^st^ generation) and most severely affected patient was the proband’s >70-year-old grandmother (patient I4). She described progressive hearing loss commencing at age 15-20, as well as two episodes of unilateral (left) acute sensorineural hearing loss over the years that partially responded to steroid treatment (Figure 2). She eventually underwent left cochlear implantation at age 50-55. In addition, she suffered from infrequent short episodes of arthritis/arthralgia over the years (Table 1). She denied recurrent episodes of fever (as well as cold-induced episodes), rash, or uveitis/keratitis. Three family members (2^nd^ generation), all in their late 30’s or early 40’s, were affected (patients II2, II4, and II6). They suffered from symmetrical bilateral mild to severe hearing loss and were hearing aid-dependent (Figure 2). In all cases, hearing impairment was initially diagnosed in their early 20’s, without additional pathognomonic systemic CAPS manifestations. One patient (II6) reported a past episode of unilateral acute hearing loss that did not respond to steroid treatment, another reported episodes of arthritis during childhood (II2), and the third sibling (II4) reported PFAPA-like episodes during childhood. Baseline inflammatory markers were mostly within the normal range in these four patients, except for patient I4 (minimally elevated CRP) and patient II2 (slightly elevated SAA) (Table ^2^).

**Figure 1.**
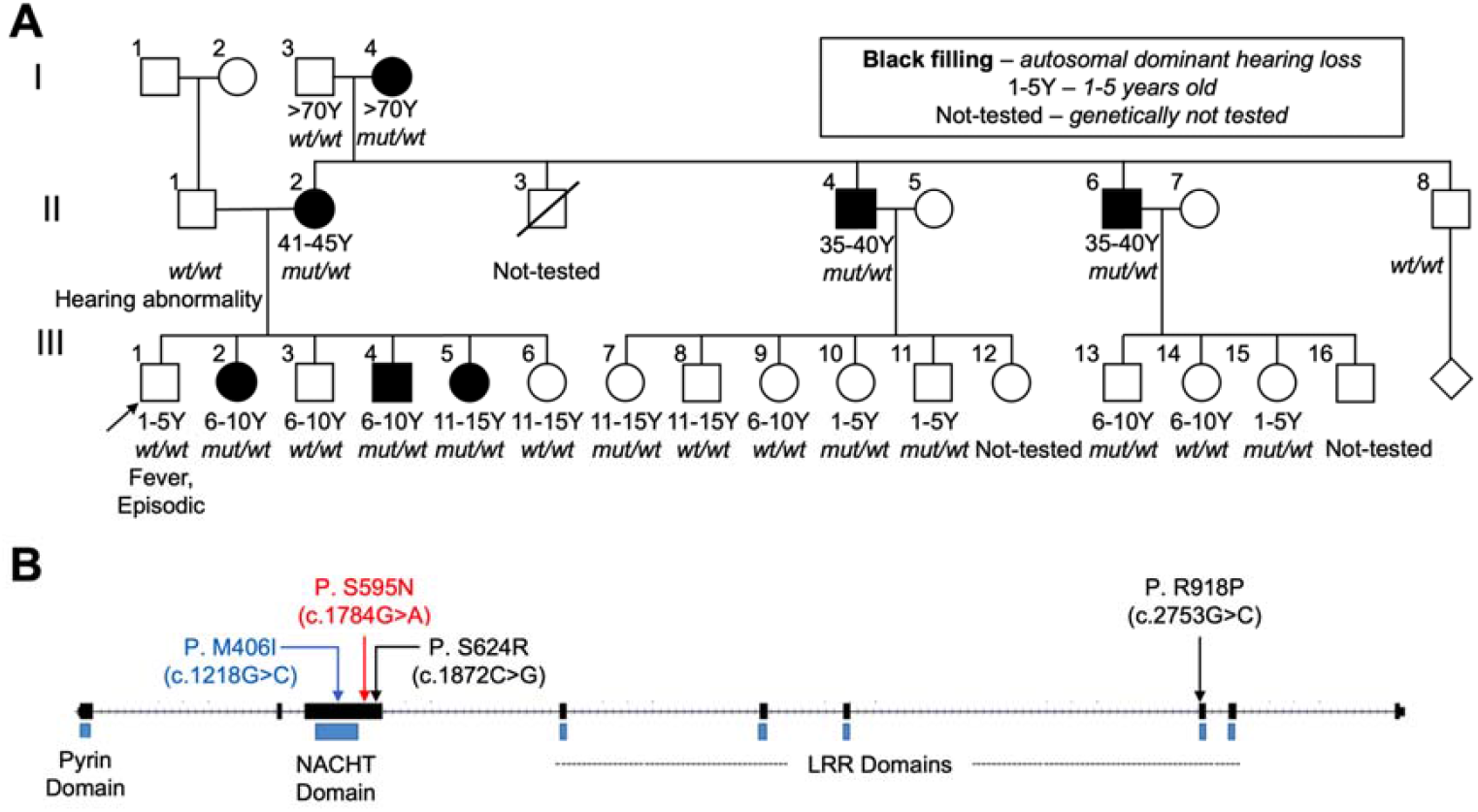
Family Pedigree and genetic information. (**A**) Pedigree of the investigated family. Subjects with autosomal dominant hearing loss are represented in green. Asymptomatic carriers and healthy family members are represented in white. Carrier status is indicated below each subject. The proband (III1) is indicated by an arrow. Pedigree was created using the Invitae family history tool (Invitae Corporation, San Francisco, California). (**B**) **Schematic view of the NLRP3 gene:** Figure adapted from UCSC genome browser, based on transcript NM_001079821.3. The figure shows only coding exons. Domain locations according to UniProt track (https://www.uniprot.org/) in UCSC genome browser (https://genome.ucsc.edu/). Variants p.Ser595Asn and p.Met406Ile, reported in this paper, are colored in red and blue, respectively. In black, previously reported variants p.Ser624Arg and p.Arg918Pro are associated with syndromic/nonsyndromic hearing loss as the primary presentation of CAPS [5,18]

**Figure 2.**
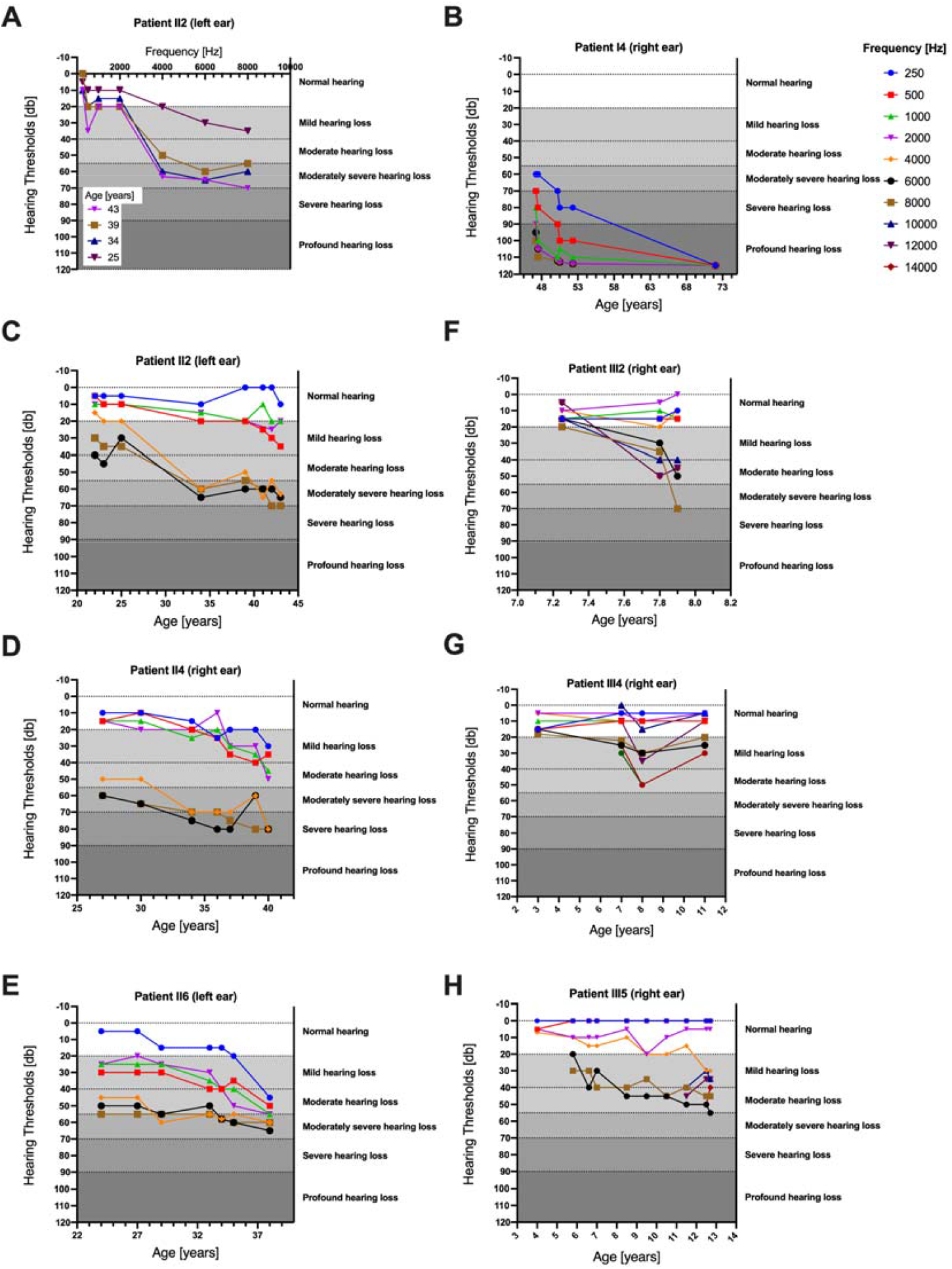
Hearing loss over time in symptomatic patients. Air conduction hearing thresholds in the better performing ear (aside from patient III4 who suffered from unilateral HL) depicting hearing deterioration over time, prior to Anakinra treatment. Patient identification number is shown on the top of each panel. (**A**) The Y axis represents hearing threshold level [dB] and the X axis tested frequency (kHz). (**B-H**) The Y axis represents hearing threshold level [dB] and the X axis age of patient (years). Each graph (in different colors) represents a different tested frequency (kHz).

**Table 1.** Clinical characteristics of family members with sensorineural hearing loss

| Subject | Age | Sex | Periodic fever | Musculoskeletal | Urticaria<br>l Rash | Eye<br>involvement | Cochlear<br>Inflammation<br>in MRI | Other*** |
| --- | --- | --- | --- | --- | --- | --- | --- | --- |
| I4 | >70 | F | No | Recurrent short<br>episodes of arthritis/<br>arthralgia | No | No | Not conducted | Rheumatic fever with<br>carditis, ITP* |
| II2 | 41-45 | F | No | Recurrent short<br>episodes of arthritis/<br>arthralgia as an<br>adolescent post<br>pharyngitis episode | No | No | No | Suspected Rheumatic fever<br>as a child (no carditis) |
| II4 | 35-40 | M | PFAPA** -<br>like resolved | No | No | No | No |  |
| II6 | 35-40 | M | No | No | No | No | No |  |
| III2 | 6-10 | F | No | No | No | No | No |  |
| III4 | 6-10 | M | PFAPA- like<br>resolved | No | No | No | No |  |
| III5 | 10-15 | F | PFAPA- like<br>resolved | No | No | No | No |  |

\* ITP- Idiopathic Thrombocytopenic Purpura \*\* PFAPA- periodic fever, aphthous stomatitis, pharyngitis, adenitis \*\*\* Other autoimmune phenomena

In contrast to the above 1^st^ and 2^nd^ generation carriers, who all suffered from prolonged and severe SNHL, most of the 3^rd^ generation carriers (15 or younger) had normal hearing function (Supplemental Figure 1). Out of the nine known 3^rd^ generation carriers, only three children suffered from hearing loss (patients III2, III4, and III5) (Figure 2 and Supplemental Figure 1C-D). Two suffered bilateral progressive SNHL; one, who initially presented with normal hearing function, was diagnosed with acute unilateral sensorineural hearing deterioration during the study period (patients III4, III5, and III2, respectively). It should be emphasized that all three children were initially diagnosed through annual auditory tests conducted considering the family history rather than due to any complaint of hearing loss at the time. No other typical CAPS signs or symptoms were present. However, both patients III4 and III5 (proband’s siblings) reported PFAPA-like episodes that resolved in the first grade. Inflammatory markers were within normal range except for minimally elevated CRP in patient III4, and mildly elevated SAA in patient III2. It should be noted that during the initial asymptomatic period, SAA levels were within normal limits; they increased in parallel to the rapid deterioration in her hearing function.

High-resolution MRI of the temporal bones were performed on all affected family members excluding the grandmother (due to technical constraints), with no signs of cochlear inflammation in the late gadolinium enhancement phase.

### Earlier detection of SNHL in ultra-high frequencies auditory evaluation

All but one affected subject suffered from progressive bilateral SNHL with a down-sloping audiometric configuration and gradual deterioration over the years (Figure 2). The SNHL pattern varied among patients, though most suffered symmetric hearing loss (Supplemental Figure 1B). Two of these subjects (patients I4 and II6) also reported past episodes of unilateral acute SNHL, which had been treated with oral steroids. One of them reported partial improvement (I4). Over the course of the study, patient III2, a seven-year-old female who initially presented with normal hearing function, suffered from acute sensorineural hearing deterioration in her right ear that solely involved the high-ultrahigh ranges (Figure 2). She was diagnosed in her routine semiannual checkup which included these frequency ranges. Patient I4 suffered from bilateral hearing deterioration over the years (Figure 2) until the left ear reached profound SNHL and required cochlear implantation. Age-eligible carriers (three symptomatic and two asymptomatic children ages 6-12) ultra-high frequencies (8KHz–16KHz) were also assessed. In all cases, the hearing deterioration was observed in the high and ultrahigh frequencies earlier than at the low frequencies (Figure 2). Pure-tone audiometry did not reveal a significant air-bone gap in any of the affected individuals.

### Identification of a novel NLRP3 variant in all patients

Patient II6 from family 1 was the first in that family to undergo molecular analysis using the Otoscope Genetic Hearing Loss next-generation sequencing (NGS) gene panel (The University of Iowa, Iowa City, Iowa), including 133 genes related to hearing loss. A heterozygous novel variant of unknown significance NM_001079821.3:c.1784G>A p.Ser595Asn (also known as NM_001079821:c.1790G>A, p.Ser597Asn) was reported on exon 5 (coding exon 3) of the NLRP3 gene (Figure 1B). It was predicted to result in a missense substitution. The genomic position does not correspond with a known domain in the NLRP3 protein (Figure 1B). However, a pathogenic variant at the same amino acid position (1783A>G p.Ser595Gly) related to CINCA syndrome was reported [11,12].

In the unrelated Family 2, patient 2.1 was diagnosed with Muckle-Wells Syndrome, based on a classical clinical presentation and genetic results. An auto-inflammatory syndromes panel (Invitae Corporation, San Francisco, California) revealed a heterozygous likely pathogenic variant c.1218G>C, p.Met406Ile in NLRP3 (NM_001079821.3) and a heterozygous variant of uncertain clinical significance in NCF2. Following parental segregation testing the variant in NLRP3 was found to be de-novo. The NLRP3 variant is located in the NACHT domain (exon 5).

It should be noted that in family 1a the father (II1) of the proband suffered from early-onset hearing loss (non-SNHL). The hearing loss gene panel did not reveal variants in his NLRP3 gene.

### NM_001079821:c.1790G>A variant carriers present functional NLRP3 inflammasome hyper activation *ex-vivo*

In order to assess whether NM_001079821:c.1790G>A, p.Ser597Asn NLRP3 variant results in inflammasome hyperactivity, we measured the levels of IL-1β secretion of peripheral-blood mononuclear cells (PBMCs) isolated from our patients, in response to priming signal alone (LPS or LPS+CaCl_2_) and in comparison to healthy controls. For technical reasons (patient recruitment to the clinic), we conducted several independent cycles of the experiment. In each cycle, we isolated PBMCs from both NLRP3 variation carriers (c.1790G>A, p.Ser597Asn) and healthy generation-matched family members as controls. Baseline IL-1β secretion levels (none treated) were comparable among all subjects. Stimulation with LPS or LPS+CaCl2, resulted in higher IL-1β secretion from NLRP3 variation carriers PBMCs compared to healthy subjects (Figure 3). The specificity of these results to the NLRP3 inflammasome activation was confirmed by using the specific NLRP3 inhibitor, MCC950, or the specific caspase-1 inhibitor, VX-765. The addition of these inhibitors to PBMCs prior to their priming by LPS+CaCl2 resulted in the reduction of elevated IL-1β secretion in all subjects except of one (Figure 3B, C). The control, Muckle-Wells pediatric patient with a known pathogenic variant in NLRP3 (NM_004895.4 c.1224G>C (p.Met408Ile), showed similar results (Figure 3D). Finally, these results were further statistically confirmed by pooling the three independent experiments and calculating the fold of change (FC) of IL-1β secretion between each treatment to the non-treated PBMCs (Figure 3E).

**Figure 3.**
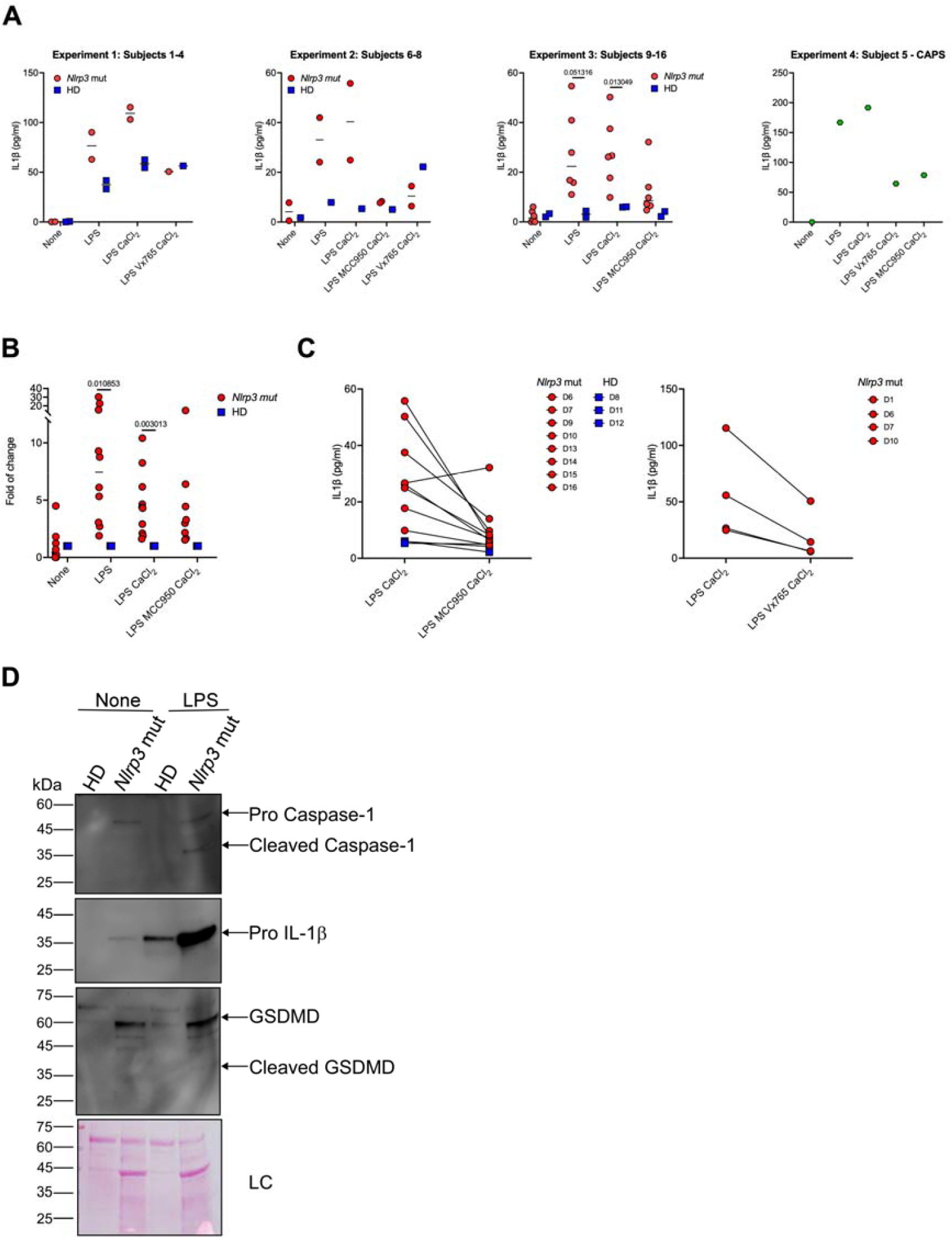
Ex-vivo functional assessment of NLRP3 activity. Approximately 10^5 PBMCs were seeded into 96-wells plate (D1-D4) or 10^6 PBMCs were seeded into 24-wells plate (D5-D16). Indicated wells were primed with lipopolysaccharides (LPS) (1 µg/mL) 3 hours prior to the experiment. Where indicated, MCC950 (2 μM) or Vx765 (25 µM) were added 30 minutes prior to the addition of CaCl_2_ (1 mM). **(A)** Cell supernatants from each experiment described above were collected 3 hours after CaCl_2_ addition or 6 hours after LPS addition. IL-1β secretion was measured using commercial ELISA kit. **(B)** Fold of change (FC) of IL-1β secreted from NLRP3 variant carriers PBMCs compared to internal healthy control of the same experiment. **(C)** Change in IL-1β secretion before and after MCC950 or Vx756 addition. **(D)** Caspase-1, gasdermin D (GSDMD), and IL-1β were detected in PBMC lysate by immunoblotting. Statistical comparisons in **(B)** between the NLRP3 variant carriers and healthy controls, were performed using multiple unpaired t-tests, *P*<0.05. In **(D)** arrows denote the expected.

To complement IL-1β secretion measurement, we analyzed the expression and cleavage of several NLRP3 inflammasome activation hallmarks: caspase-1, GSDMD, and IL-1β using immunoblot and specific antibodies. As can be seen in Figure 3G, pro-caspase-1, pro-IL-1β, and full-length GSDMD were detected in the PBMCs of NLRP3 variation carrier (subject I4) prior to LPS stimulation whilst remaining at undetected levels in the healthy control (subject no.2). Furthermore, in agreement with IL-1β secretion phenotype, GSDMD and caspase-1 in NLRP3 variation carrier PBMCs were detected in their activated (cleaved) state (Figure 3D). Overall, these *ex-vivo* functional results suggest that c.1790G>A, p.Ser597Asn is a gain-of-function mutation that results in a constitutively unique semi-activated basal state of the NLRP3 inflammasome.

### Substantial hearing improvement in response to IL-1R antagonist treatment (Anakinra)

The clinical presentation, family segregation of the novel variant, and evidence of inflammasome hyper-activation in the above *ex-vivo* functional studies suggest that the novel variant 001079821:c.1790G>A,p.Ser597Asn is a pathogenic gain-of-function mutation-causing atypical CAPS phenotype, according to the Eurofever classification criteria [13]. Thus, we decided to treat the affected patients with an interleukin-1 receptor antagonist (IL-1Ra) drug, which blocks IL-1β activity. Therapy with IL-1Ra (Anakinra) was consequently administered at an initial dose of 3 mg/kg (up to 100 mg) daily. Clinical follow-up included clinical and hearing assessment as well as the measurement of inflammatory markers 1-3 months post-treatment initiation (Tables 2, 3). Audiometry reassessment was conducted one and three months after treatment initiation. The three affected children showed rapid and near-complete reversal of hearing impairment, and returned to near-normal hearing function within 1-3 months (Figure 4). Among the three 2^nd^ generation subjects, we observed minimal asymmetrical improvement in hearing function (Figure 5). Although the grandmother (patient I4) with the cochlear implant did not report any improvement in hearing function, she did report a resolution of arthralgia within two weeks of treatment initiation. In addition, all her initially slightly elevated inflammatory markers returned to normal within one month of treatment (Table 2.).

**Table 2.** Inflammatory markers levels pre- and post- Anakinra

|  | <u>Prior to Anakinra Treatment</u> |  |  |  | <u>After 3 months of Anakinra Treatment</u> |  |  |  |
| --- | --- | --- | --- | --- | --- | --- | --- | --- |
| Patient | WBC | ESR | CRP | SAA | WBC | ESR | CRP | SAA |
| I4 | 7.2 | 36 | 0.84 | 5.3 | 4.42 | 8 | 0.16 | 2.8 |
| II2 | 7.54 | 28 (old) | 0.56 | 22.7 | 5 | 24 | <0.05 | 4.7 |
| II4 | 6.9 | 15 | 0.25 | 6.2 | 7.12 | 10* | 0.09 | X |
| II6 | 7 | 12 | 0.52 | 5.5 | 7 | 5 | 0.07 | X |
| III2 | 11.13 | 19 | 0.26 | 4-12.6<br>** | 7.88* | 21* | 0.43* | X |
| III4 | 10.4 | 14 | 1.19 | <0.82 | 5.2 | 13 | 0.6 | 3 |
| III5 | 6.6 | 15 | 0.64 | 3.3 | 5.71 | 5 | <0.05 | 1 |

\* 1 month after Anakinra initiation \*\* Original levels of SAA were undetectable. However, SAA levels were measured and elevated during sudden hearing loss episode. White blood cells (WBC) K/micl, ESR- mm/1hr, CRP- C reactive protein mg/dL, SAA - Serum Amyloid A mg/l (range 0.0-6.4)

**Table 3.**
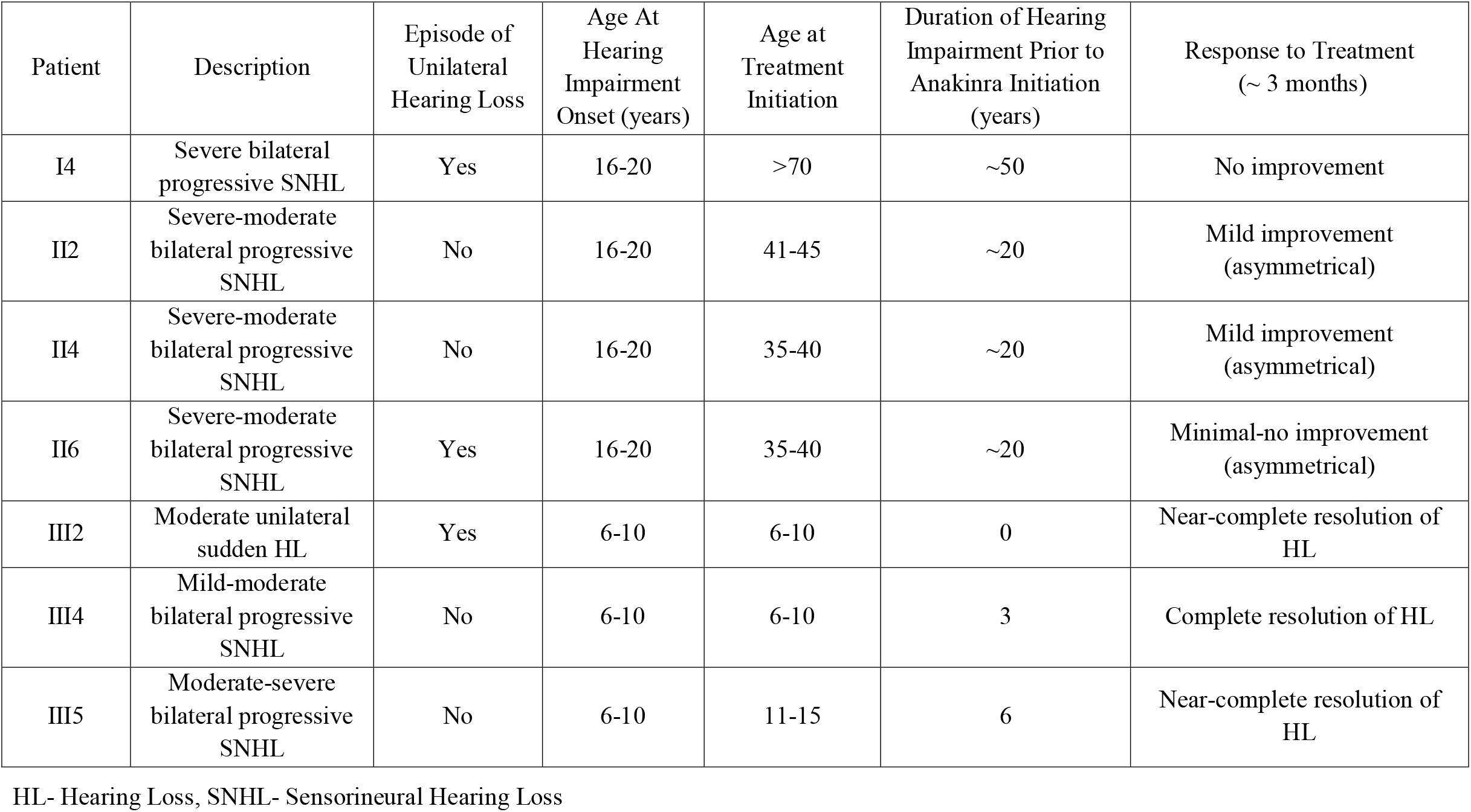
SNHL features and response to Anakinra treatment

**Figure 4.**
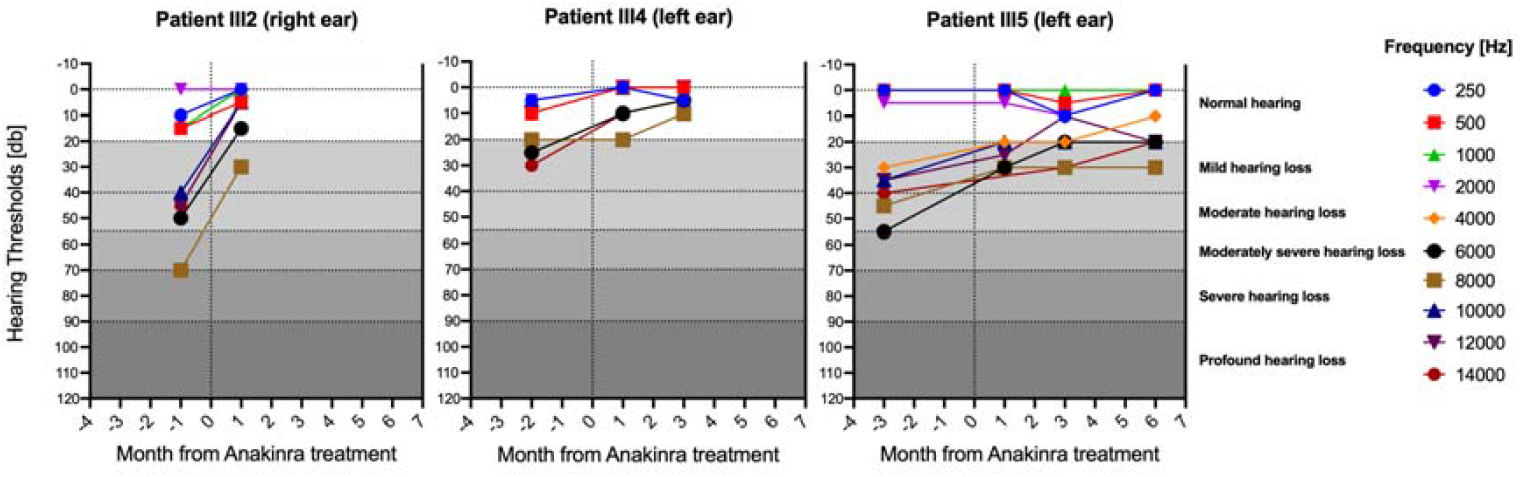
Anakinra restores hearing loss in pediatric symptomatic patients. Air conduction hearing thresholds depicting dynamics in hearing function over time, in response to Anakinra treatment. Patient identification number is shown on the top of each panel. The Y axis represents hearing threshold level [dB] and the X axis time from treatment initiation (months). Each graph (in different colors) represents a different tested frequency (kHz).

**Figure 5.**
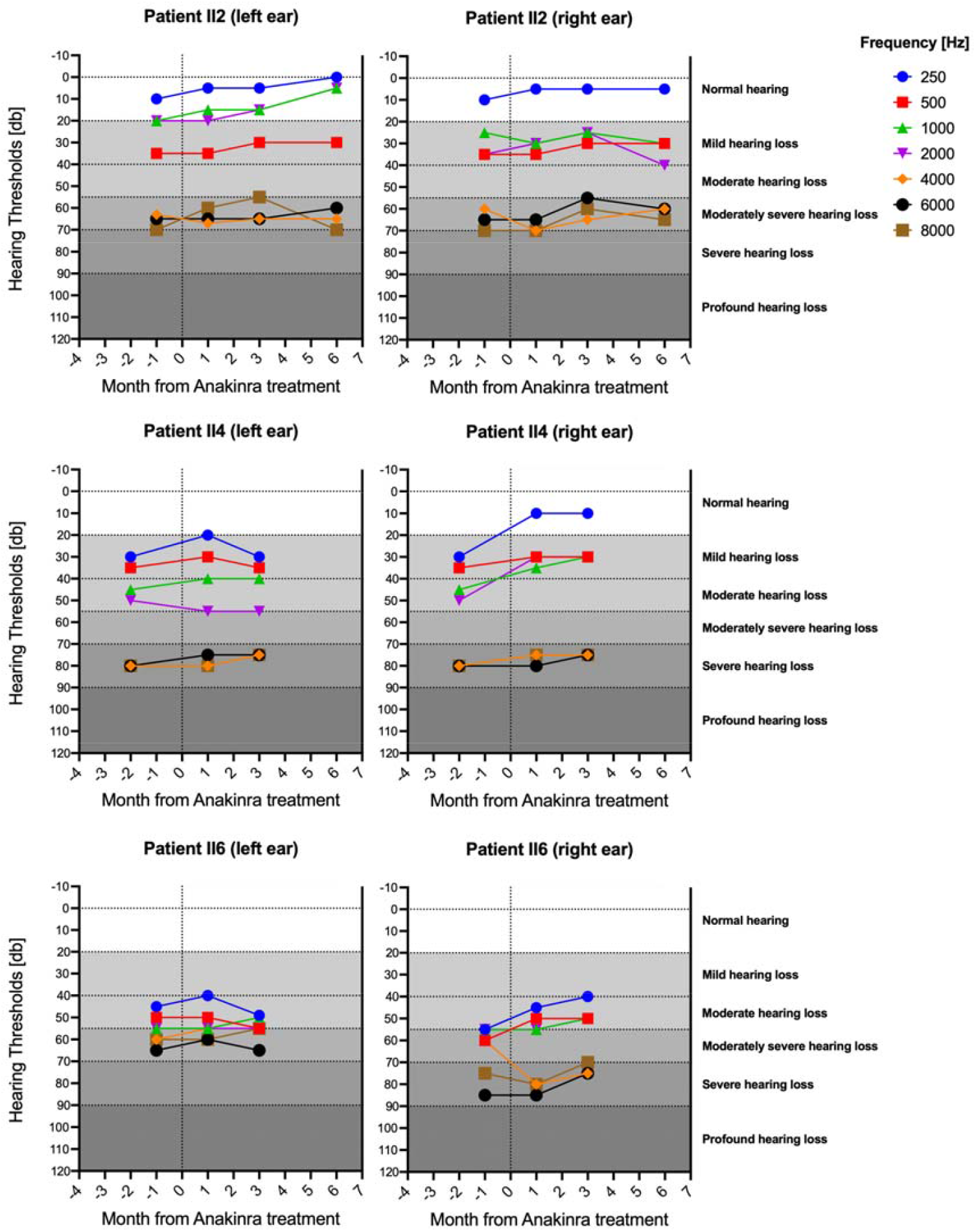
Anakinra slightly improves hearing loss in adult symptomatic patients. Air conduction hearing thresholds depicting dynamics in hearing function over time, in response to Anakinra treatment. Patient identification number is shown on the top of each panel. The Y axis represents hearing threshold level [dB] and the X axis time from treatment initiation (months). Each graph (in different colors) represents a different tested frequency (kHz).

No severe adverse reactions to Anakinra were reported.

## Discussion

Gain-of-function mutations in the NLRP3 gene led to hyper-activation of the NLRP3 inflammasome, resulting in excessive secretion of inflammatory cytokines such as IL-1β and IL-18. Clinically these mutations cause a spectrum of autosomal-dominant systemic autoinflammatory diseases known as CAPS/NLRP3-AIDs. In addition to the three classical clinical subtypes of CAPS (NOMID, MWS, and FCAS), there have been a few reports of NLRP3 missense variants associated with progressive sensorineural hearing loss as the primary/sole clinical manifestation [5,6,14].

In this study, we presented family members spanning three generations with a novel missense variant in the NLRP3 gene of unknown significance (NM 001079821:c.1790G>A, p.Ser597Asn), with clinical features inconsistent with classical CAPS (i.e., past history of PFAPA, sparse episodes of isolated fever, and nonspecific past history of arthralgia/arthritis). Even though they did not meet the classification criteria for CAPS [13], the familial genotype-phenotype segregation and potentially treatable hearing loss (autosomal dominant progressive/rapid/mixed SNHL) led us to explore the suspected diagnosis of atypical CAPS. Thus, in the hope of establishing the suspected diagnosis and enabling prompt initiation of appropriate anti-IL-1 therapy, we conducted further clinical, imaging, and functional inflammasome assessment.

Autosomal dominant progressive SNHL initially affected the high and ultrahigh frequency range (as seen in pediatric patients III2 and III4) and progressed to severe impairment involving lower frequencies also in adult patients (Figure 2). In addition to the typical CAPS-associated bilateral progressive deterioration [15], some subjects suffered from an atypical pattern of acute unilateral hearing loss (I4 and II6) (Figure 2). In the course of this study, subject III2, a 5-10-year-old female who initially presented normal hearing function, suffered from unilateral rapid sensorineural hearing deterioration that involved only the high-ultrahigh ranges (Figure 2). It should be notes that she had no auditory complaint s and was diagnosed in her routine semiannual checkup which included these frequency ranges. To the best of our knowledge, this is the first report of isolated acute hearing loss among CAPS patients. Furthermore, the prompt initiation of Anakinra treatment in this patient, which led to nearly complete resolution of SNHL (Figure 4), highlights the importance of both the routine auditory evaluation of the high-ultrahigh ranges and rapid anti-IL-1 intervention treatment.

Interestingly, although the affected adults reported hearing impairment onset in their early 20’s, early symptomatic and asymptomatic changes in the high and ultrahigh-frequency ranges were detected among the affected children as early as five years of age (Figure 2F-H). This could be explained either by phenotypic progression over generations, increased awareness of hearing impairment among family members, or merely thanks to early detection in semiannual hearing function tests starting from infancy that specifically included the high and ultrahigh-frequency ranges. Nevertheless, regardless of the reason for this early detection, this further supports the importance of adding these frequency ranges to the regular auditory evaluation of these patients.

In addition to hearing impairment, further clinical evaluation of known carriers revealed a high prevalence of PFAPA in childhood among 4/13 carriers. This is an interesting finding, as although the pathogenesis of PFAPA syndrome is unknown, it has previously been linked to inflammasome hyper-activation [16].

Surprisingly, a magnetic resonance imaging (MRI) of the temporal bone did not reveal any signs of cochlear inflammation among affected family members. However, it should be noted that the grandmother (I4) did not complete an MRI due to cochlear implant incompatibility. In addition, as recently described [17], cochlear MRI may be normal in some CAPS patients with SNHL and is considered a positive predictive factor for clinical response to Anakinra treatment.

As described, in order to further establish the diagnosis, we conducted functional assessment of the NLRP3 inflammasome activity in PBMCs of carriers vs. healthy family members. As expected, secreted IL-1β levels and cleavage of GSDMD and caspase-1 were substantially and significantly higher among carriers vs. healthy controls following priming without the secondary and specific activation of the NLRP3 inflammasome. These results are consistent with the dogma that in cases of gain-of-function mutations in NLR as NLRP3, IL-1β secretion can be induced by priming signals such as LPS. [18,19]

The response to Anakinra treatment was dramatic, especially among the 3^rd^ generation pediatric patients. The three affected children showed near-complete reversal of hearing impairment and returned to near normal hearing function within 1-3 months (Figures 4, 6). Not surprisingly, the grandmother (I4), who presented the most severe and prolonged symptoms, did not report improvement in hearing function, probably as a result of irreversible cochlear damage on the right and the presence of the cochlear implant on the left. However, she did report a complete resolution of arthralgia within two weeks, and inflammatory markers normalized within a month of treatment. 2^nd^ generation patients (II2, II4, and II6) who suffered from severe and prolonged hearing impairment (threshold below 60dB in some frequencies) (Figures 2C-E) still gained an overall slight improvement in hearing function (Figures 5-6). Although this could be a result of irreversible cochlear damage after years of untreated cochlear inflammation, it could also be explained by the relatively low initial Anakinra dosage (100 ng per day). For this reason, we are currently considering increasing the daily dosage. In line with our observation, a recent study [17] categorized seventeen families diagnosed with either CAPS or atypical CAPS according to the severity of hearing impairment, MRI findings/absence of evidence of cochlear inflammation, and evidence of NLRP3 hyper-activation (according to an in vitro LPS stimulation test). They suggested that a hearing function threshold below 60dB prior to Anakinra (Anti-IL-1) treatment and cochlear enhancement on brain MRI are related to poor audiological prognosis and responsiveness to Anakinra therapy.

**Figure 6.**
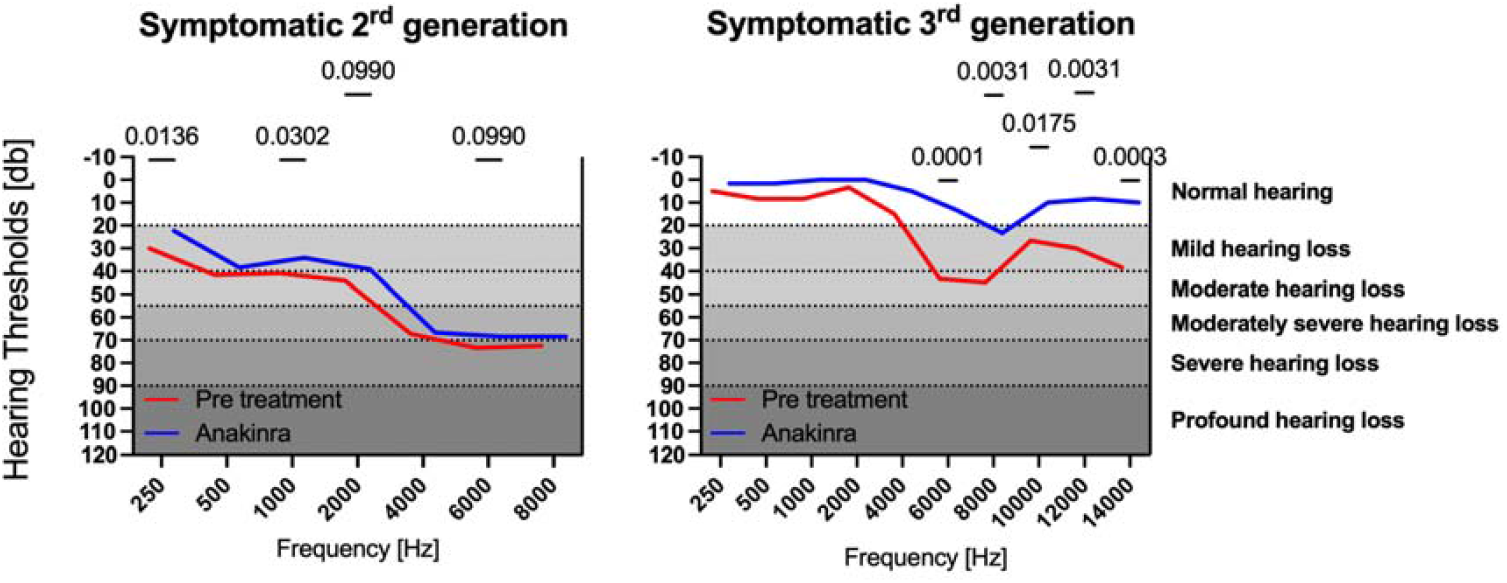
Comparison of hearing loss Anakinra restoration in adults and pediatric symptomatic patients. Air conduction hearing thresholds representing dynamics of hearing function one month after initiation of Anakinra treatment in 2^nd^ (N=3, both ears were tested) and 3^rd^ (N=3, one ear, the affected ear was tested) generation patients (left and right panel, respectively). Generation specified on the top of each panel. The Y axis represents the hearing threshold level [dB], and the X axis tests frequency (kHz). Averages of pre-treatment is represented in red and post-treatment in blue. In the Statistical comparisons between the pre- and post-treatment were performed using two-way ANOVA (Fisher’s LSD test), Significant (*P*<0.05) is shown.

In summary, our findings highlight the crucial role of early diagnosis and treatment with an anti-IL-1 agent in reversing cochlear damage. Furthermore, our results suggest that high and ultrahigh frequency ranges should be included in the auditory assessment to enable early detection of subclinical SNHL. Finally, functional inflammasome assessment can establish the diagnosis in inconclusive cases.

## Material and methods

### Patients

A family of Jewish Ashkenazi descent initially presented with autosomal dominant bilateral SNHL with a progressive/rapid/mixed deterioration pattern. The hearing impairment initially affected high/ultra-high frequencies as an isolated presentation or was accompanied by nonspecific systemic signs and symptoms inconsistent with typical CAPS (i.e., sporadic episodes of fever, arthritis/arthralgia, or PFAPA (periodic fever, aphthous stomatitis, pharyngitis, adenitis)). All SNHL family members were carriers of a novel missense mutation in NLRP3 of unknown significance and did not fulfill CAPS classification criteria [13] (Figure 1A, Table 1). We assessed ten known carriers spanning three generations who were found to be carriers of the NLRP3 variant, seven with impaired and three with normal hearing function. Age at study initiation ranged from 2-70 years. The control group consisted of five healthy family members who were matched by age group. Our study group also included one unrelated 5-10-year-old patient (P 2.1) who presented with classical signs and symptoms of Muckle-Wells and is a carrier of a known heterozygous pathogenic variant (NM_004895.4) in NLRP3. This patient served as a positive control for inflammasome hyperactivation in the study.

### Clinical and genetic evaluation

Affected family members underwent clinical evaluation by a clinical rheumatologist, otorhinolaryngologist, and a clinical geneticist. Medical records and past audiometry tests were reviewed. Clinical evaluation and peripheral blood inflammatory markers were obtained for all symptomatic patients before and after interleukin-1 receptor antagonist (IL-1Ra) (anakinra) treatment initiation. Pure-tone audiometry was performed per test eligibility (age-dependent) on all known carriers, of whom seven were symptomatic (I4, II2, II4, II6, III2, III4, III5). The ultra-high frequency range (8KHz – 16KHz) was also evaluated in patients III2, III4, III5, III7, and III9. Follow-up audiometry was conducted on all treated patients.

High-resolution magnetic resonance imaging (MRI) scans of the temporal bones were performed with a Philips Ingenia 3 Tesla system (Philips Healthcare). The scans included an axial heavily T2 weighted sequence (DRIVE) and a T1 weighted sequence. Following the injection of Gadolinium-based contrast media, a 3D sequence (3D FLAIR or 3D T1 Dixon) was performed.

### *Ex-vivo* functional assessment of NLRP3 inflammasome activity

Peripheral blood samples were drawn into an EDTA tube from a total of 15 subjects (11 novel variant carriers, one Muckle-Wells patient, and three age-matched healthy family members as controls). PBMCs were immediately isolated (up to two hours from collection) as done previously [20]. See Supplementary Data for further details. Approximately 10^5^ PBMCs were seeded in each well of the 96 well plate and LPS was added to indicate wells for three hours. Where indicated, caspase-1 inhibitor, Vx765, or NLRP3 inhibitor, MCC950, were added 30 minutes prior to CaCl_2_ addition. Cell supernatants and cell lysates were collected three hours later for further analysis. Supernatants were analyzed for IL-1β secretion using ELISA kit (Invitrogen Waltham, Massachusetts, USA). Cell lysates were analyzed for cleaved human-IL-1β, cleaved caspase-1, and cleaved GSDMD using immunoblot and specific antibodies. See further details in supplementary data.

### Statistics

Statistical comparisons in the *ex-vivo* experiments between the different treatments were performed using multiple unpaired t-tests. A comparison of hearing before and after Anakinra treatment was preform using RM two-way ANOVA. P values <0.05 were considered statistically significant.

### Data analysis

Data were calculated using GraphPad Prism 9, and details can be found in the figure legends.

## Data Availability

All data produced in the present study are available upon reasonable request to the authors

## Data availability statement

The original contributions presented in the study are included in the article/Supplementary Material. Further inquiries can be directed to the corresponding authors.

## Ethics statement

All experiments in this study were reviewed and approved by the ethics committee (RMC0941-20) and performed according to the Declaration of Helsinki. Prospective and retrospective reviews of medical history, laboratory and imaging results, rheumatologic evaluations, and laboratory tests were performed after informed consent. Written informed consent was obtained from adult subjects and from both parents of minor subjects.

## Author contributions

BM initiated and designed the study concept and coordinated the multidisciplinary assessment of patients; BM, CH, and GM designed the experiments; BM, and CH performed experiments; SM, RE, and HO performed the clinical hearing evaluation; KL performed the brain image analysis; KN, SE, and ON coordinated and conducted clinical-genetic assessments; AG, DR, LG, BM, and HL, performed the clinical evaluation and management; BM, wrote the original draft; All authors wrote and edited the manuscript. HL, and GM contributed to the study conception and design and supervised the work.

## Acknowledgments

First and foremost, we would like to extend our thanks and appreciation to all family members who participated in this study. We would also like to express gratitude to our colleagues Ivona Aksentijevich, Kalpana Manthiram, and Amanda Ombrelo from the National Institute of Health; and Ziv Erlich from Tel Aviv University for their assistance throughout.

## Contribution to the field statement

Cryopyrin-associated periodic syndromes (CAPS) are a spectrum of rare auto-inflammatory diseases caused by gain-of-function mutations in the NLRP3 gene, resulting in inflammasome hyper-activation and subsequent uncontrolled release of Interleukin-1β (IL-1β). Many patients with CAPS develop progressive sensorineural hearing loss (SNHL) due to cochlear auto-inflammation, which in rare cases may be the sole or main manifestation. In this study, we establish the suspected diagnosis of CAPS in a family presenting autosomal dominant SNHL with progressive/acute deterioration and a novel missense variant in the NLRP3 gene of unknown significance (NM_001079821:c.1790G>A, p.Ser597Asn), by conducting *ex-vivo* functional assessment of the NLRP3 inflammasome. Subsequent administration of anti-IL-1 therapy resulted in a substantial clinical improvement especially among pediatric patients who exhibited near resolution of hearing impairment within a few months of treatment. Our findings highlight the crucial role of early diagnosis and treatment with an anti-IL-1 agent in reversing cochlear damage. Furthermore, our results suggest that high and ultrahigh frequency ranges need to be included in the auditory assessment to enable early detection of subclinical SNHL. Finally, incorporating functional inflammasome assessment as part of the clinical evaluation could establish the diagnosis in inconclusive cases.

## Supplemental material

### Peripheral blood mononuclear cells (PBMCs) isolation

PBMCs were isolated from peripheral venous blood samples freshly drawn from subjects 1-16. Briefly, 10-30 ml of peripheral blood were collected and loaded on Histopaque-1077 (Sigma Aldrich 10771). Cells were centrifuged for 30 min at 400 g at 24°C. Cells from the interphase were collected and washed with PBS. Samples of the cells were then stained for flow cytometry to determine the percentage and concentration of monocytes, before they were seeded in a 96-wells plate or a 24-wells plate at a final concentration of 10^6 monocytes/ml, in DMEM supplemented with 10% heat-inactivated fetal bovine serum (FBS) (Gibco, 10270106). After 18 h, the wells were washed three times to remove non-adherent cells and medium was replaced with DMEM supplemented with 1% FBS. Cells were treated with 1ug/mL ultrapure LPS. After 3 hours of LPS treatment, 2 ug/mL MCC950 or 25 µM Vx765 were added to selected wells for 30 minutes prior to adding 1mM CaCl2 for 3 hours. After 3 hours cell supernatants and cell lysates were collected for further analysis by ELISA or immunoblot, respectively. Experiments were performed according to the guidelines of the Institute’s Helsinki Ethics Committee.

### Immunoblot analysis of proteins

Cells were collected and centrifuged for five minutes at 400xg in order to separate them from the supernatant. Next, the cells were lysed by using RIPA buffer in the presence of protease inhibitors at 4°C for 15 min. Lysed cells were loaded onto any kD gradient Criterion TGX-Free precast gels (Bio-Rad, Hercules, California, USA). Proteins were transferred onto a nitrocellulose membrane (Bio-Rad), and Ponceau S staining was performed routinely to evaluate the loading accuracy. Membranes were blocked with 5% (w/v) skim milk in TBS for 1–2 h, and then probed overnight with primary antibodies (all diluted 1:1000, unless noted otherwise): pro and cleaved human-IL-1 β (R&D, AF-201-NA), pro and cleaved human caspase-1 (Merck, 06-503-I), and pro and cleaved human-GSDMD (abcam, ab210070). Relevant horseradish peroxidase-conjugated secondary antibodies were applied for at least 1 hour. Membranes were washed between antibody incubations four times in TBS containing 0.1% (v/v) Tween 20 (TBST). Antibodies were diluted in TBST containing 5% skim milk. Immunoblots were developed using an ECL kit (Bio-Rad) in an ODYSSEY Fc (Li-COR, Lincoln, Nebraska USA) equipped with Image Lab software. All images were cropped for presentation. Full-size images will be presented upon request.

### Reagents

Unless stated otherwise, all cell culture reagents were purchased from Biological Industries, Beit-Haemek, Israel. Lipopolysaccharides (LPS) of Escherichia coli O111:B4 were purchased from Sigma-Aldrich (#L3024, Munich, Germany). Vx765 and MCC950 and ELISA kit were purchased from Invitrogen (Waltham, Massachusetts, USA). HRP-conjugated secondary antibodies were purchased from Jackson ImmunoResearch Labs (West Grove, PA, USA).

## Supplemental Figures

**Supplemental Figure 1.**
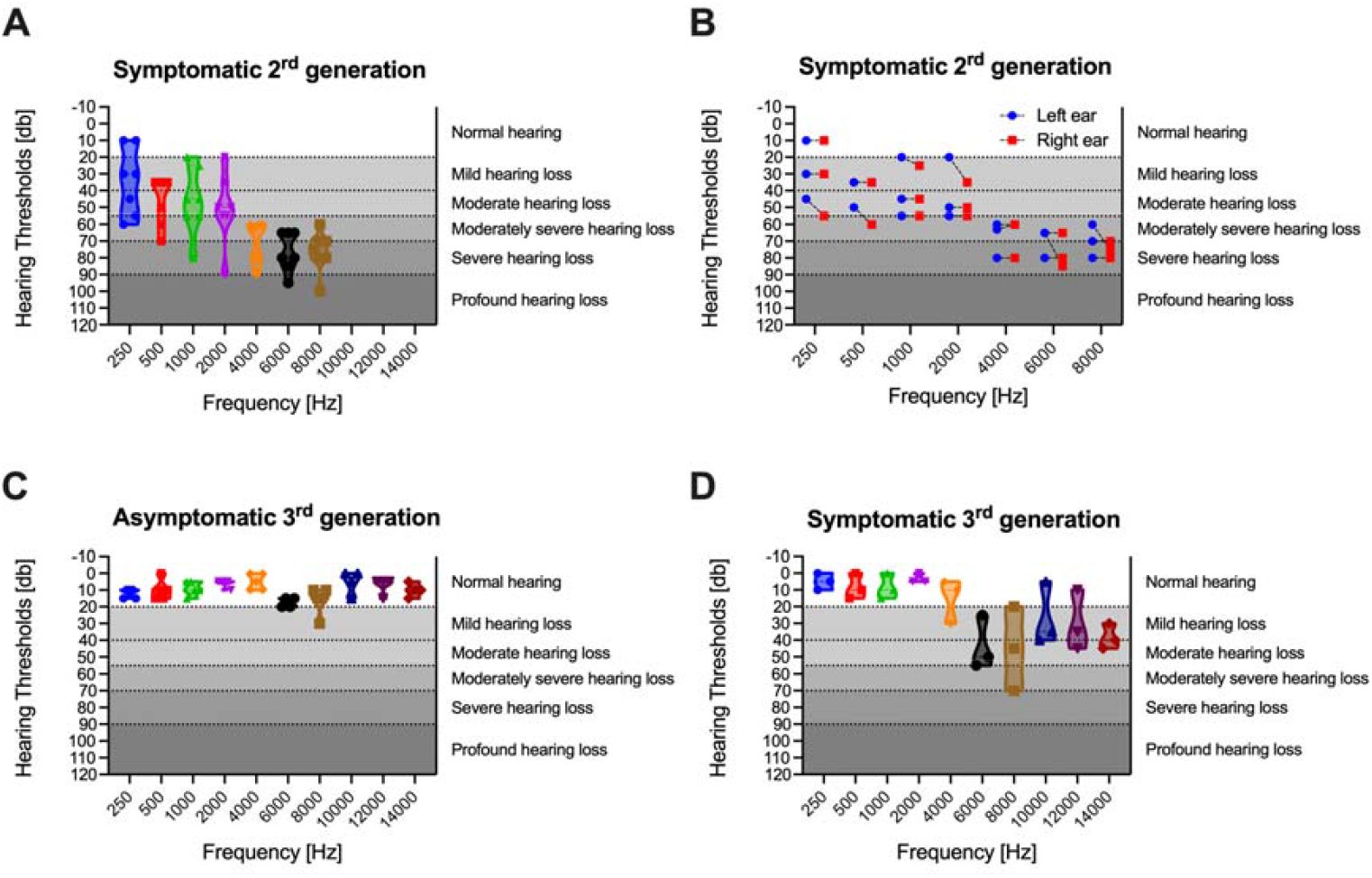
Hearing loss 2^nd^ and 3^rd^ generation patients. Pooled same-generation auditory phenotypes based on generation and hearing loss status are stated on the top of each panel. Although hearing loss severity per tested frequency varied among patients, overall, the high-ultrahigh hearing range is most severely affected (panels A, D).

